# Carer reported experiences: supporting someone with a rare disease

**DOI:** 10.1101/2020.07.10.20150581

**Authors:** Julie McMullan, Ashleen L. Crowe, Kirsten Downes, Helen McAneney, Amy Jayne McKnight

**Affiliations:** Centre for Public Health, School of Medicine Dentistry and Biomedical Sciences, Institute of Clinical Science Block A, Grosvenor Road, Belfast, BT12 6BA

## Abstract

**Background:** The uniqueness and complexity of rare diseases, together with a perceived lack of understanding from health professionals, can make caring for someone with a rare disease extremely challenging. Carers are often forced to become ‘expert’ patients alongside people they care for. Due to the demands placed upon carers it is essential that appropriate support is available for them to ensure they can continue to carry out this vital role. This exploratory study researched challenges carers face when looking after someone with a rare disease and identify how they could be better supported in this role.

**Methods:** To be eligible to participate, respondents had to be adults caring for someone with a rare disease. Mixed methods were used including an online survey (n=57 respondents) where carers highlighted a need for better support specific to their mental health needs, liaising with health and social care professionals, financial, communication, training and respite options. During a facilitated workshop (n=32 attendees) discussions focused on challenges carers face as well as priorities to better support carer needs in the future.

**Results:** While carers reported several positive aspects of their caring role, the majority of comments highlighted challenges such as sub-optimal interactions with healthcare professionals, insufficient (or absent) emotional, psychological and social support, lack of financial support, and lack of awareness of existing support services.

**Conclusion:** It is important that strategies are put in place to ensure that carers take the time they need to care for themselves and raise awareness of available support options for carers of people with a rare disease(s) from health and social care providers, charities, or support groups.

**Strengths and limitations of this study:**

- This study provides insight into current challenges, and some requested solutions, based on reported experiences from carers of people with a rare disease in the UK and Ireland.
- The use of an online survey promoted flexibility and accessibility for person’s unable to attend the in-person workshop
- The facilitated workshop enabled carers to network, discussing challenges and potential solutions with their peers.
- Responses in this exploratory study are based upon people who were self-motivated to participate.

## Introduction

Rare diseases are collectively common and affect a significant proportion of the population^1^, representing a major public health issue. It is estimated that over 450 million persons worldwide (6 – 8 % world population) have a rare disease^2^, with more than four million persons affected across the UK. Support for people living and working with rare diseases is currently inadequate^3, 4^ with challenges including a lack of understanding by health and social care professionals, delayed diagnosis, and difficulties gaining optimal treatment^5-7^.

Due to a low prevalence and lack of expertise for many of the >6000 rare diseases, patients are often forced to become knowledgeable about their own disease state^8^. They become ‘expert patients’ alongside their carer as they seek an empowering and collaborative approach with their clinicians^9^. This is a shift from the traditional patient-doctor relationship, with each party revising their role and expectations, which presents many challenges^10, 11^. Caring for someone with a rare disease, both formally and informally, can be an extremely demanding role requiring intense and unique care tailored to each individual’s specific needs^12^. There may be significant merit in peer support from networking with other carers experiencing similar circumstances themselves^13^.

The impact of caring for someone with a rare disease can be seen in many areas of an individual’s life including psychologically, economically, physically and logistically^14^. The importance of good mental and physical health for the carer is vital to ensure they are able to sustain the essential role which they provide for the individual they care for^15^. Neglecting to look after themselves can have devastating consequences on each carer’s own health, which directly impacts the person they care for^16^.

There is no consensus scale to capture experiences from formal and informal carers of people with rare diseases across all age groups. For example, the Carer Experience Scale (CES) is an index measure of the caring experience focused on economic evaluations^17^. The ASCOT-carer scale is a self-report instrument designed to measure social care-related quality of life for family-friend unpaid carers^18^, while the FAMCARE-2 scale has been used to measure carers’ perceived satisfaction with palliative service provision^19^. The exploratory approach undertaken in this study aimed to gain an understanding of carer reported experiences derived specifically from persons caring for someone with a rare disease.

## Methods

A qualitative approach was chosen^20^ with experiences reported by carers supporting individuals with a rare disease(s) sought *via* a survey and facilitated workshop. All participants provided written, informed consent. Ethical approval was provided by the Faculty of Medicine, Life and Health Sciences, Queen’s University of Belfast (QUB) research ethics committee (MHLS 19_08).

### Patient and Public Involvement

Patients and public were involved in the design of this survey and helped to promote participation. The project aim was a direct result of previous research within the QUB Rare Disease Team which highlighted the need for research focusing solely on carers. It became apparent that carers are often overlooked in rare disease research and yet they play such a vital role in the life of someone with a rare disease. As part of the dissemination of results we plan to develop a leaflet to be given to carers at time of diagnosis. We have sought input from carers and rare disease patients as to how this should be formatted and what information should be included.

### Online survey

An online survey was hosted by <u>SmartSurvey</u>^21^ 6th November 2019 – 31^st^ January 2020 with five sections and 43 questions (supplementary file 1). Questions were designed using an iterative approach with the <u>Northern Ireland Rare Disease Partnership</u>^22^ and were focused on caring responsibilities, interactions with health care and support services, networking, communication and future improvements. Closed and open-ended questions were employed, with the option to save the survey and complete at a later date.

Survey respondents identified as caring for an individual with a rare disease, aged over 18 years old. The survey was promoted via the NIRDP website^22^ and was shared on their Twitter (@NI_RDP; 1168 followers) and Facebook (@NIRDPNews; 1013 members) sites. Queen’s University Belfast also promoted the survey via their websites and Twitter pages. An A5 flyer (supplementary file 2) was developed to promote the survey displaying a QR code and a link to the survey and distributed locally across QUB campus and libraries. The survey was also promoted at two rare disease events held locally in Newtownards (December 2019) and Bangor (January 2020), Northern Ireland. Charts and crosstabs were generated using SmartSurvey and MSExcel to gain frequencies and percentages. The qualitative data from open questions was analysed thematically.

### Facilitated workshop

A workshop for carers of people with a rare disease was organised in association with the NIRDP^22^ on 16^th^ January 2020 in Bangor Carnegie Library, Northern Ireland. The workshop was advertised in a similar manner as for the survey. The workshop was an open event, anyone was welcome to attend. It lasted approximately two hours. The event was primarily an opportunity for carers to network with one another as well as with multiple carer focused organisations based in Northern Ireland who were in attendance highlighting their services (supplementary file 3).

An independently facilitated discussion at the workshop was conducted with all attendees and using small groups. Notes were taken during the discussion and analysed by 2 researchers (AM & JM). Three key themes were explored during discussions, focusing on each for approximately 20 minutes:

1. ‘positive aspects of the caring role’
2. identifying and prioritising the challenges carers face and to consider how these problems might be overcome.
3. what good support may look like and priorities to improve resources for carers across Northern Ireland.

## Results

### Survey

#### Respondent characteristics

Fifty-seven respondents participated in the survey, with 56 identifying the person they care for as a family member (Table 1). Respondents resided in Northern Ireland, Great Britain, or the Republic of Ireland. 80.7% respondents identified as caring for the individual in a voluntary/unpaid capacity, while others identified specifically as being a parent, or wife, and so may also fall under the voluntary role although not explicitly stated; one respondent identified as a paid carer. 87.7% of respondents live with the person they care for, with 35.7% caring for children. A table showing the demographics of respondents is presented below.

**Table 1:** Demographics of survey respondents

| Demographics of Respondents |  | % | Frequency<br>(n=57) |
| --- | --- | --- | --- |
| Gender | Male | 15.8 | 9 |
|  | Female | 84.2 | 48 |
| Age, years | 25-34 | 14.0 | 8 |
|  | 35-44 | 24.6 | 14 |
|  | 45-54 | 49.1 | 28 |
|  | 55+ | 12.3 | 7 |
| Country of residence | Northern Ireland | 96.5 | 55 |
|  | Great Britain | 1.8 | 1 |
|  | Republic of Ireland | 1.8 | 1 |
| Number of individuals currently cared for* | 1 | 77.2 | 44 |
|  | 2 | 10.5 | 6 |
|  | 3 | 10.5 | 6 |
|  | 4 | 1.8 | 1 |
| Relationship to the person cared for | Family | 98.2 | 56 |
|  | Friend | 0 | 0 |
|  | Other (started as client, now a friend) | 1.8 | 1 |
| Age of the person(s) cared for* | Less than 18 | 35.7 | 20 |
|  | 18-24 | 16.1 | 9 |
|  | 25-34 | 12.5 | 7 |
|  | 35-44 | 7.1 | 4 |
|  | 45-54 | 14.3 | 8 |
|  | 55+ | 19.6 | 11 |
|  | Prefer not to say | 1.8 | 1 |
| What capacity they care for the person | Voluntary / unpaid | 80.7 | 46 |
|  | Paid-employed | 0 | 0 |
|  | Self-employed | 7 | 4 |
|  | Other | 12.3 | 7 |
| Live with the individual cared for | Yes | 87.7 | 50 |
|  | No | 12.3 | 7 |
| Geographic setting | Urban | 47.4 | 27 |
|  | Rural | 52.6 | 30 |

\*please note that the number of individuals cared for is 78, but that a person may have 2 or more persons cared for in separate age categories.

Half of carers (∼50%) look after an individual full time/24 hrs a day. The majority of respondents (86.0%, n=49) do not have any respite care available. Of the 14% (n=8) who do have respite care available 75% (n=6) of them have used this respite facility. Prior to caring for the individual with the rare disease 54.4% (n=31) of the respondents had not heard of a rare disease, and 75% (n=42) of the respondents had not had any interactions with someone with a rare disease.

### Training for carers

Only 14.3% (n=8) of the respondents had received formal training to assist with their caring for the individual with a rare disease. Completed training included medication administration, hoisting, tube feeds, wheelchairs, oxygen administration, first aid, manual handling, safeguarding, working with people with learning difficulties, maintaining airways, Multi Agency Public Protection Arrangements (MAPPA) training. When asked if they would like to receive formal training as a carer 25.5% (n=14) said they would like to receive in person training, 25.5% (n=14) said they would like a workshop, 7.3% (n=4) said yes to it being delivered online at a specific time, and 38.2% (n=21) said yes to it being delivered online but accessible in their own time. Of the respondents only 20% (n=11) said they would not like to receive formal training as a carer.

### Understanding of genetics and multi-omics

More than ^4^/_5_ rare diseases have a genetic cause ^23^, yet only 3.5% (n=2) of respondents identified as having a confident understanding of genetics. Approximately one-quarter of respondents would like to know more about genetics (26.3%, n=15) and multi-omics (24.6%, n=14).

### Mental Health

All respondents (100%, n=57) stated that caring for a person with a rare disease impacted their mental health. Only 12.3% (n=7) of respondents stated that they had received psychosocial support in relation to their role as a carer. When asked what they had received or what services they had accessed one individual described a brief conversation with a GP, another spoke of self-referred to counselling. Others had used cognitive behaviour therapy, a pain management, mental health course, landmark worldwide (https://www.landmarkworldwide.com/) and one person said *“Always have nurses or doctors to call if worried about anything 24/7”*.

### Medical appointments and health care professionals (HCPs)

In relation to medical appointments, dealing specifically with rare disease, 39.2% (n=16) described them as being a positive experience and 69.8% (n=37) as a negative experience (see figure 1a and b). The majority of respondents (80.7%, n=46) did not feel that medical professionals had sufficient knowledge to look after those with a rare disease (figure 1b).

### Networking

Half of respondents (54.4%, n=31) were involved in one or more support groups, with a similar number having contacted a rare disease group (52.7%, n=29), and significantly less a carers group (12.3%, n=7). Reasons for contact included seeking peer support/feeling isolated; information about policy; experiences from other families; about the disease after getting diagnosis; what services and help are available; offering help; to seek out medical experts on the field; to seek diagnosis and to raise funds. Of the respondents who had contacted a charity/support group, 70.6% (n=24) said they had their need met. Respondents reported communicating with other carers most frequently *via* internet forums (45.9%, n=17) and in person (29.7%, n=11). Many stated that communicating with other carers reduces isolation and keeps them in a positive mind set. When asked if networking with different sets of people would be of interest the majority of respondents answered yes (supplementary file 4). Only 1 respondent thought that sufficient support is available for those who care for someone with a rare disease with several suggestions made to improve support (figures 2 and 3).

Less than half of respondents (40.4%, n=23) had attended a rare disease conference/event. Of those who did attend, 18 (78%) found it very useful, 4 (17%) said it could have been better, and 1 (4%) said they would not go back. Many carers were unable to attend such events due to lack of time, lack of respite care, and insufficient resources including financial. The challenges experience by carers and priorities for carers are summarised in Figures 4a and b.

### Facilitated workshop/discussion

The first topic for discussion focused on the positive aspects of being a carer. Carers reported feeling a sense of pride; being able to help the person to enjoy life; giving life purpose and fulfilment; and gaining an increased medical knowledge. This was followed by discussion of the biggest challenges for carers. Interactions with HCPs was highlighted as a major hurdle and carers stressed their need to be taken seriously in such settings. Ideas such as a feedback system for HCPs and raising awareness of the importance of treating individuals with rare diseases, and those caring for them, with dignity and respect were suggested. Insufficient emotional, psychological and social support were highlighted with an advice helpline suggested as an approach to help.

Financial support was discussed with many carers explaining that carers allowance is not sufficient and many are not considered eligible to receive it. There was a consensus that carers should be recognised and adequately paid for their role.

Carers requested information on services such as respite and where to access specific equipment. They reported regular feelings of guilt due to the lack of attention they give siblings because of the demands put upon them by their caring role for a child. Several participants suggested that respite services would be welcomed to enable taking other family members for a short break. The majority explained that current respite services are generally inadequate for their needs and suggested having respite carers who understand individual medication needs; good communication would help improve this service. Many improvements carers suggested included online training, websites which link relevant resources, organisations, and psychosocial support.

## Discussion

Research focusing on rare disease often does not include carers yet given the unique role they have in the person’s care it seems crucial that their views are heard^24-27^. Our research emphasises challenges faced by carers supporting individuals with a rare disease and highlights areas where support is lacking.

### More Support Required (including finances, training, mental health, and respite)

The majority of attendees highlighted their financial struggles as carers for individuals with a rare disease. This was in both priorities of what would help most and in biggest challenges. For example, there are significant additional costs associated with raising a child with a disability^28^, with 38% of parents having to reduce their paid employment hours or leave work all together^29^. Many adult rare disease patients employ carers under the ‘direct payments’ scheme; spouses and children living in the same household are not eligible to receive ‘direct payments’ funding for their caring roles^30^, despite the frequent need for 24/7 care. Although there is sometimes a small amount of funding available from the government as a carer’s allowance, it is not nearly enough to cover costs of medical treatment and travel^13 31^.

Unanimously carers experienced insufficient support, which is a common issue internationally^13, 29, 32 33, 34^. Several respondents perceived there was more support available for more common diseases, with so little resources available for rare diseases. Provision of additional support, or improved signposting towards existing resources would minimise some of the strain described by carers. A particular area highlighted in both the survey and facilitated discussions was additional support requested for carers and siblings of children with a rare disease(s). Lagae (2019)^33^ described how 91% of carers feel caring makes family life difficult, stating that 46% of siblings had missed a leisure opportunity in the past 2 weeks due to parental caring responsibilities.

A request for accessible respite was raised many times in this study, which complements international reports where respite support is not adequately provided and would be highly valued to help with day to day management of their loved one’s symptoms^5, 35-38^.Practical support is very important to minimise excess strain on carers, particularly when carers feel their friends and family cannot empathise with their situation^39^.

Formal training for carers is lacking. Parents of those with a rare disease have expressed that they would value receiving training to assist them in their caring needs^40^ as would young carers^41^. Training for carers would enable improved, safer care and may facilitate better relationships with HCPs. Providing training to primary carers for people with complex care needs may result in cost-savings for healthcare providers by minimising adverse events and unplanned hospital admissions. More than a third of the respondents preferred training delivered online to flexibly work with their caring commitments.

All (100%) of carers reported that caring has affected their mental health. Limited support for emotional/psychological/social wellbeing was also raised many times during the discussions. Additional practical issues reported in the literature included the limited help they receive for things like transportation, shopping and chores^42^. With recent findings showing 84% of carers in the UK feel more stress due to their caring role^43^.it is a common problem which needs to be addressed. A survey by Pelentsov 2016 reported 37% of carers for someone with a rare disease were being treated for depression, 41% for anxiety, and 10% for other mental illnesses^29^. There is a clear need for improved psychosocial support for carers of people with a rare disease(s).

### Awareness and Information

Awareness and information were mentioned repeatedly by respondents seeking ‘*more information’*. A lack of awareness was highlighted for the impact that caring for someone with a rare disease has on the life of each carer and their family. Internationally, carers of those with a rare disease have asked for educational resources for friends/family^44^, ^37^ Providing accessible educational resources for families and the community as a whole would be a practical step to help raise awareness of the role of carers and common impacts when caring for someone with a rare disease. A discussion area in this project that we did not find featured in the literature was a need for a lobbying body for carers of those with rare disease.

A lack of appropriate information and communication for rare diseases has been highlighted with individuals seeking more information about rare disease diagnoses, implications of long-term care plans, and community support teams to provide clarity for carers and help them feel more supported ^35^, ^45^, ^44^, ^8^. Carers often have incomparable personal knowledge of how a rare disease affects the person they support. In spite of this knowledge, carers are rarely provided with sufficient practical or medical information to assist them in their role as a carer^44^, ^36^. Information particularly sought in this study focused on information about diagnosis, treatments, support, and finances. Focused on information at the time of diagnosis, “*More carers information given at diagnosis. This is the single most important thing*.*”* was not something commonly reported in the literature, although more timely diagnosis and better access to diagnosis were referred to several times ^5, 44, 45^. Although this may not be the single most important to thing to all carers, information around the time of diagnosis would support carers to make choices. Ultimately, carers of those with a rare disease are living in an information vacuum where so many of us take the access and ease to ‘common’ information for granted.

### The Experts (Healthcare professionals and carers)

Sub-optimal interactions with HCPs have been highlighted repeatedly in studies across the globe; our workshop and survey, predominantly derived from Northern Ireland (NI), showed no exceptions and indeed poorer experiences reported than observed internationally. For example, 81% of carers felt their HCP did not have enough knowledge to treat their rare disease, compared to 54% of parents reporting their HCP lacked adequate knowledge^29^. And this is in fact opposite to an Australian study which reported 73% of carers felt their GP had adequate knowledge of their rare disease^13^. Many carers were dismayed feeling left to figure out the disease themselves due to the lack of understanding by HCPs, which echoes other findings^12^. A lack of knowledge by some HCPs can have detrimental effects, including delaying diagnosis^46^, which is already a stressful event^5^. Where relevant, carers being recognised as experts in a rare disease is necessary, but not often the case^47, 48^. Carers are keen to have equality with patients when it comes to healthcare services “*The same respect for my needs as that which the disabled person is entitled to”*. Carers being recognised and treated equally would streamline the of the patient, where the health care system relies on carers to carry out so much of the day to day functions necessary for a patient’s wellbeing. Collaboration with healthcare professionals is mentioned in the literature as something of value and which has a positive impact on carers^47-49^. Carers were adamant they would like to receive more respect as carers and experience improved communication between HCPs with both patients and carers, particularly with respect to co-developing care plans.

### Social Isolation

Social isolation was frequently reported by carers with many experiencing no free time for themselves while caring for individuals up to 24 hours a day. 86% of participants reported that there is no available respite for the person they care for, which is inevitably going to affect the social life of the carer and in turn their mental health. Similarly Pelentsov and colleagues observed 58% of carers losing friends since they stared caring for someone with a rare disease^29^ while 80% of carers said caring made socialising difficult^33^. Online support groups have been documented in the literature to help with mental health effects of caring^32^. Of our survey respondents, 70% of those currently in communication with other carers used the internet or social media to communicate. The flexibility in regards location of an online platform makes it easy for carers who cannot arrange alternative care for the person with a rare disease whom they support. When the respondents were asked about networking the predominant result was that there was a preference of networking with healthcare professionals and other carers rather than charity workers or patients. Increased capacity for networking would facilitate connection and improve information dissemination and sharing.

### Strengths and Limitations

This research study is based on the experience of carers of people with a rare disease in the UK and Ireland. It highlights the challenges that they are facing and allows for them to put forward their own solutions and suggestions for support, understanding, and information. Using an online survey was deemed the most efficient way to reach a large number of carers in a short time period. The hope was to get a range of views from a wide geographical spread such as from both urban and rural settings, as well as between age ranges of carers. Carers often find attending events a challenge due to their caring responsibilities, so making the survey available online made it more accessible^50^. Online surveys offer flexibility as they can be completed at a time suitable to the individual and have a function to enable it to be completed in stages^51^, saving progress as they work through the questions. They also allow anonymity and the opportunity to skip questions that they are not comfortable answering^51^.

Given that no appropriate carer scale survey could be found in the literature, the questions of this survey were designed specifically for this study using an iterative approach, with input from the NIRDP^22^. The survey focused on the challenges carers’ face, the support they receive and how they feel they could be helped.

The workshop participants may have been a biased sample as all were self-motivated to attend, and the survey respondents were self-motivated to complete the survey. Though both the carers of children and adults with a rare disease are included here, and the survey was open to paid and voluntary carers.

The workshop also provided a valuable opportunity for those caring for people with a rare disease to network, which is valuable to carers^49^, and to meet representatives from voluntary organisations. The carers surveyed were based predominately in NI, which provides an important local evidence base for policy and practice implications.

## Conclusion

Carers of those with a rare disease are asking for better access to psychosocial support, better financial provision for their substantive role, improved access to helpful information to give clarity for the future, training to assist them in their role, and options for respite care. Improved interactions with HCP’s and primary carers that encompasses both understanding and recognition of carer’s crucial role in the life of whom they care for. A carer experience scale is required for carers of people with a rare disease. While many themes are common across all carers, some are unique to rare disease carers, for example the difficulties experienced at appointments with health care professionals and the lack of awareness and information surrounding rare diseases. Future surveys for ‘carers’ should include a question asking if they care for someone with a rare disease so that their unique needs can be identified. The time taken for carers to provide such detailed and valuable responses to this study demonstrates how much those caring for someone with a rare disease need improvement both for their own quality of life, and to enable them to enrich the life of who they care for by having better provisions to facilitate that care.

## Data Availability

The datasets generated and/or analysed during the current study are not publicly available due as they contain potentially identifiable information but are
available from the corresponding author on reasonable request.

## Declarations

### Competing Interests

The authors have no competing interests to declare. AJM is a former board member and HM an existing board member of the NIRDP.

AJM and JM conceived of the project, designed the survey with input from patients, healthcare professionals and voluntary groups. JM and AC drafted the manuscript. JM and KD collected the survey data. AC, JM, and KD analysed the data. AJM and HM reviewed the manuscript. All authors contributed to data interpretation, manuscript revision and agreed the final version for submission.

## Acknowledgements

We thank all individuals and groups who participated in this study. We also thank NIRDP (www.nirdp.org.uk) and QUB (https://www.qub.ac.uk/research-centres/CentreforPublicHealth/) for helping promote the study.

## Legends for figures

Figure 1a and 1b: Experience of healthcare appointments and healthcare professionals.

Figure 2: Responses when asked what could be done to help.

Figure 3: What would you like to see improved regarding support for carers?

Figure 4a: Biggest challenges when caring for someone with a rare disease.

Figure 4b: Themes of priorities that would help most in caring for someone with a rare disease.

